# Literature review of Diabetes noninvasive screening tools for previously undiagnosed. Scoping review

**DOI:** 10.1101/2023.12.19.23300192

**Authors:** Tesfaye Haile

## Abstract

**Background:** Diabetes is a severe chronic disease that needs due attention. Worldwide distribution of the disease is increasing in alarming trend marked by its pandemic nature. Numerous people are disabled and dead due to the complexity of the disease and its management. A recent CDC report revealed that in the USA, 37.3 million people have Diabetes (11.3% of the US population). Ninety-six million people aged 18 years and older have prediabetes (38.0% of the adult US population). Worldwide, the number of people with Diabetes rose from 108 million in 1980 to 422 million in 2014. WHO on its sustainable development goal (SDG) report made clear the existing diabetic patient evaluation, intervention, and monitoring method does not adequately address the problem. Paradoxically, the ambitious goal of the American Diabetes Association is: “A Future Without Diabetes.”

The current Diabetes screening method is invasive, provided that scarcity of resources and high cost of test kit makes it nonpractical to frequently checkup. We must proactively engage in its prevention by making screening cheap and easily accessible. So, this review is designed to answer the question: What is an available Diabetes noninvasive screening tool for undiagnosed?

**Method:** The relevant Article was searched from PubMed, MEDLINE, Embase, Web of Science, SCOPUS, and the CINHAL using a combination of the following key constructs: Diabetes screening, Diabetes noninvasive screening, Diabetes AND screening, Diabetes and (“noninvasive” or “non-invasive” or “non invasive”) and “screening tool,” Diabetes AND noninvasive AND screening AND Tool, “Diabetes noninvasive Screening Tool.” The selection process was conducted based on the PRISMA framework statement. Exclusion and inclusion criteria were applied to select the final relevant literature, after which 22 relevant studies were selected.

**Result:** The review included studies conducted from 2013 to 2023, 36% in 2019 and 2022. The included articles are from 13 countries. The data sources for the articles are surveys, databanks, random sampling, and opportunity sampling. A total of 24 key predictors were used to develop 22 types of tools. The researchers used the logistic regression (LR) method while developing most studies, and ML in a couple of them. The Authors described the Tool’s performance using the ROC curve, and the AUC is 0.65-0.93. The developed tool predictive power of the screening Prediabetes used Sensitivity, Specificity, PPV, NPV metrics and the result is encouraging.

**Conclusion:** Diabetes noninvasive Screening tests have the potential to identify prediabetes at an early stage and, thus, a more treatable disease that, as a result, saves lives. The developed tools had promising designate ability in pre-DM case findings, as shown by their ROC curve, AUC, sensitivity/specificity, and PPV/NPV. The Tool developed by these researchers looks promising for our goal of screening Diabetes by noninvasive method to answer this review research question using non-laboratory methods, which can be applied by common people regardless of the involvement of health professional skill. This review’s limitations are that the included article’s bias assessment was not performed, and non-English language articles were not included, so this could miss some pertinent tools. Finally, a systematic review, meta-analysis, and RCT on the Tool are recommended to identify any bias during the development of the Tool and possible generalizability of the best Tool for worldwide applicability.

## Introduction

Diabetes is a severe chronic disease that needs due attention. Numerous people are disabled and dead due to the complexity of the disease and its management.^1^ Worldwide distribution of the disease is increasing in alarming trend marked by its pandemic nature.^2^ The June 29, 2022, report of the Centers for Disease Control and Prevention (CDC) revealed that in the USA, 37.3 million people have Diabetes (11.3% of the US population).^1^ Ninety-six million people aged 18 years and older have prediabetes (38.0% of the adult US population).^1^ Worldwide, the number of people with Diabetes rose from 108 million in 1980 to 422 million in 2014.^2^ Thus, Prevalence has risen more rapidly worldwide.^2^

World health organization (WHO) on its sustainable goal (SDG) report made clear the existing diabetic patient evaluation, intervention, and monitoring method does not adequately address the problem.^2^ Paradoxically, the American Diabetes Association vows “A Future Without Diabetes.” So, the question is, how do we solve this paradox? One option is to focus on preventing early metabolic abnormalities and implementing routine screening.^2^ However, the current Diabetes screening method is invasive (blood drawing and test for blood glucose) provided that scarcity of resources and high cost of test kit makes it nonpractical to frequently checkups, which is a hurdle to early detection of the deviation of metabolic status from the typical standard set point. So, it is better to have a self-administered, noninvasive screening tool used frequently at a convenience. This screening tool is less dangerous, less expensive, and less time-consuming and causes less physical and psychological discomfort. Thus, this review is designed to answer the question: What is an available Diabetes noninvasive screening tool for undiagnosed? This review search is intended to look for the presence of self-administered Diabetes noninvasive screening tools for the general population use from existing studies. Therefore, the rationale for this review is to look for easy tools for self-administered noninvasive Diabetes screening to effectively act to prevent this silent killer disease by detecting its early marker of metabolic abnormality. Knowing the regular status of Diabetes vulnerability in a very accessible way can enable early detection of this metabolic abnormality and facilitate the correction and prevention endeavor.

### Aim

Searching for Diabetes noninvasive screening tools for undiagnosed.

### Objective of the study

to look for the research availability on Diabetes noninvasive screening tools by a systematic process, transparent strategy, standardized data extraction methods, and present summary of findings.

### The research question

What is available Diabetes noninvasive screening tool for undiagnosed in existing works of literature/studies?

### Methodology

After developing a search strategy together with an experienced CUNY SPH librarian, Rosemary Farrell, MA, MLS; we searched PubMed, MEDLINE, Embase, Web of Science, SCOPUS, and the CINHAL using a combination of the following key constructs: Diabetes screening; Diabetes noninvasive screening; Diabetes AND screening; Diabetes and (“non-invasive” or “non invasive” or “noninvasive”) and “screening tool”; Diabetes AND noninvasive AND screening AND Tool”; “Diabetes noninvasive Screening Tool”. These databases were chosen because they cover many scientific articles on eHealth. The selection process was conducted and presented based on the PRISMA framework flowchart. (see Figure 1) All duplicates were checked thoroughly and removed using Microsoft Excel to maintain the quality of the review. The titles and abstracts of the articles were checked closely for analysis and purification to ensure the articles’ relevance for eligibility checks. Exclusion and inclusion criteria were defined based on relevant literature, after which 22 relevant studies were selected for further review. The Inclusion and exclusion criteria for selected studies were the following.

**Figure 1.**
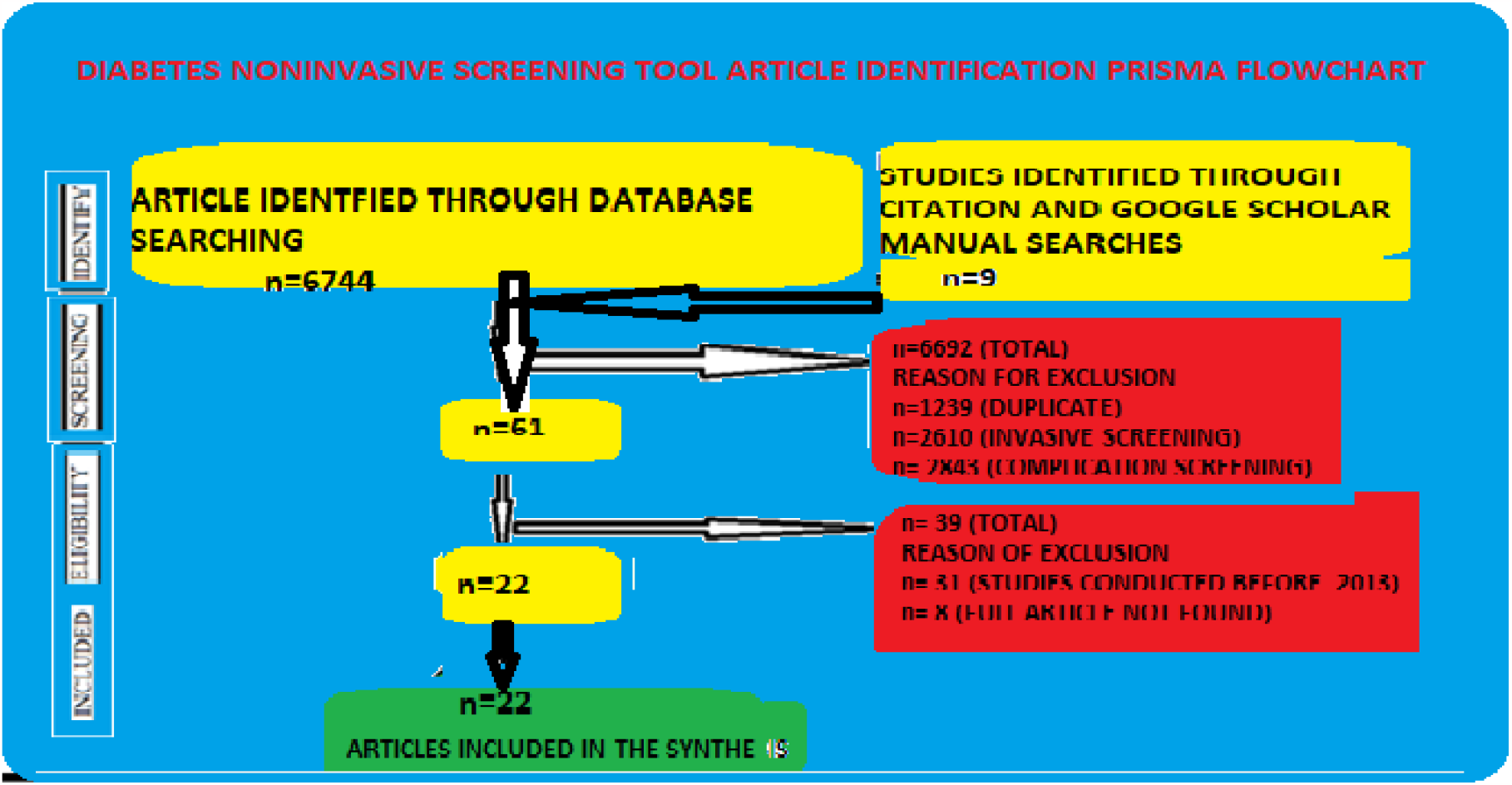
PRISMA flow chart showing the article selection process

### Inclusion criteria

The Article should be:

1. Diabetes noninvasive screening tool
2. The study published from 2013 to 2023
3. Presented in the English language
4. Full text accessible
5. The Tool is easy to be applied by common people

### Exclusion criteria

The Article should not be:

1. Diabetes Screening by Imaging Procedure
2. Diabetes screening methods need instrument and result interpretation involving health professional
3. screening of Diabetes complication

### Data Extraction and Analysis

After the articles fulfilled the eligibility criteria, they were retrieved for review. The following general characteristic data were extracted: the authors, year of publication and country of origin, the aim, methodology of the study, sample source and sample size, the Tool outcome measurement [referenced by the’ Gold standard’ (FBS, RBS, 1h PG, 2hr PG, and HbA1c)], number of the identified Key predictive findings,

Predictors Tool, Tool performance evaluation, and the tool predictive powers. The extracted Data was presented (displayed) on tables, charts, and figures for better insight acquisition and synthesized narratively, and the findings were then summarized.

## Result

A total of 6,744 articles were identified from the database searches. Six thousand six hundred ninety-two were removed due to duplicates using Microsoft Excel, and articles out of the scope of the review (invasive screening and Diabetes complication screening) were screened deeply based on titles and abstracts and then excluded. Based on eligibility criteria, 39 articles were removed—15 full articles were retrieved for further review. Nine additional studies were identified through citation and Google Scholar manual searches, with seven meeting the inclusion criteria. Finally, 22 studies were included in the review process (See Figure 1).

From the 22 studies included in this review, a total of 24 key risk predictor were identified for 22 Tool development. Table 1 summarizes the included studies’ characteristics and prediction tools.

**Table 1:**
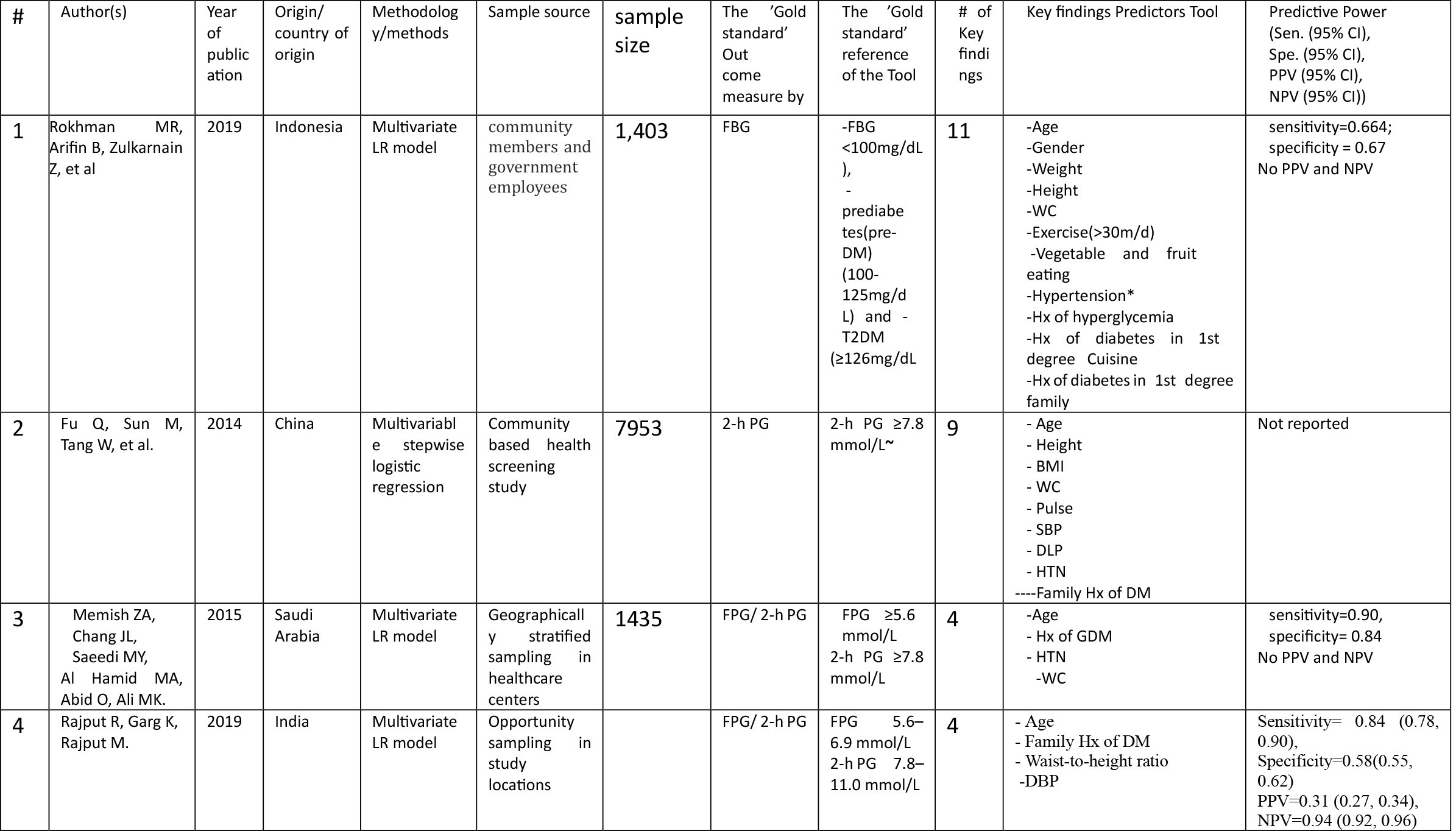
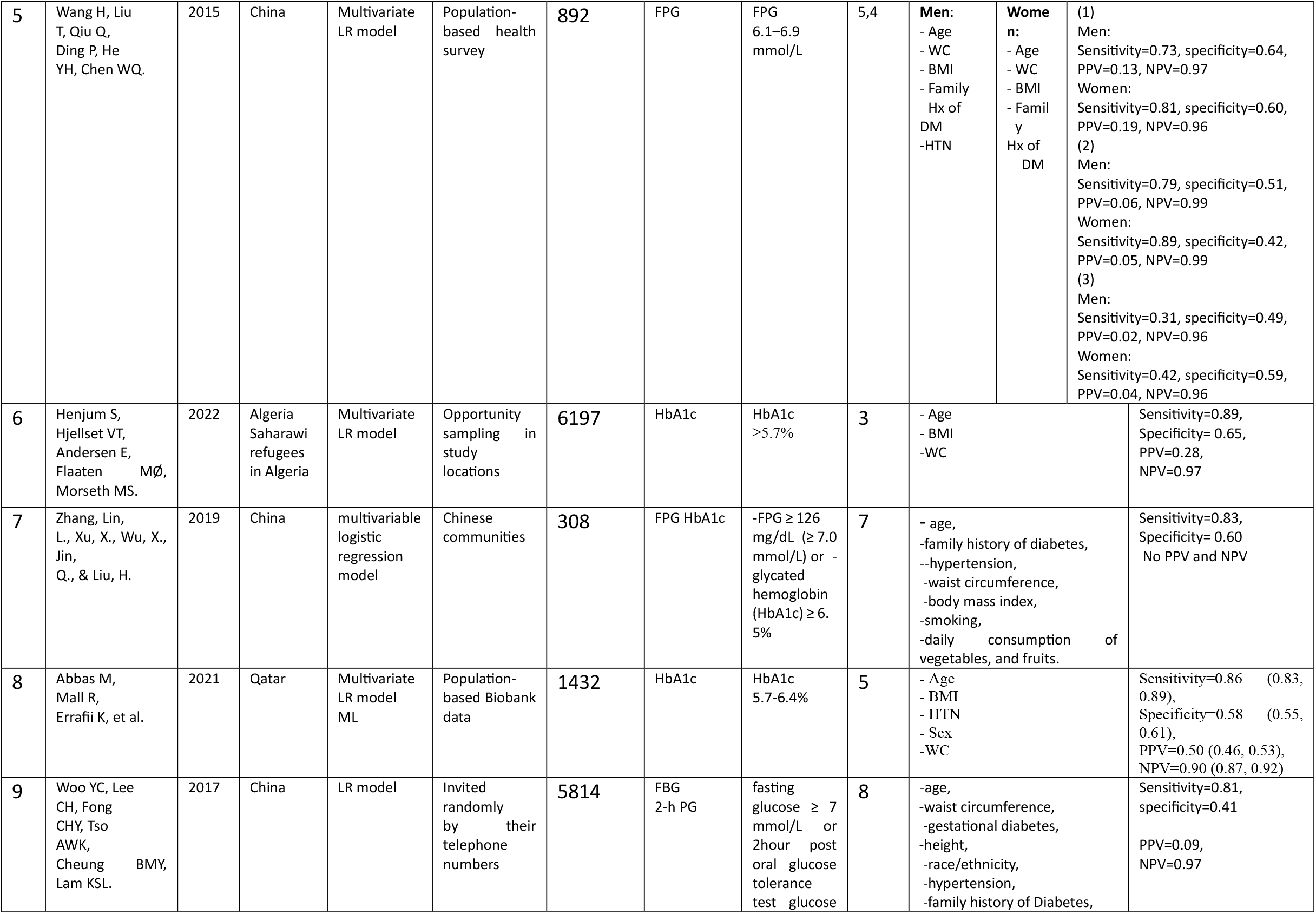
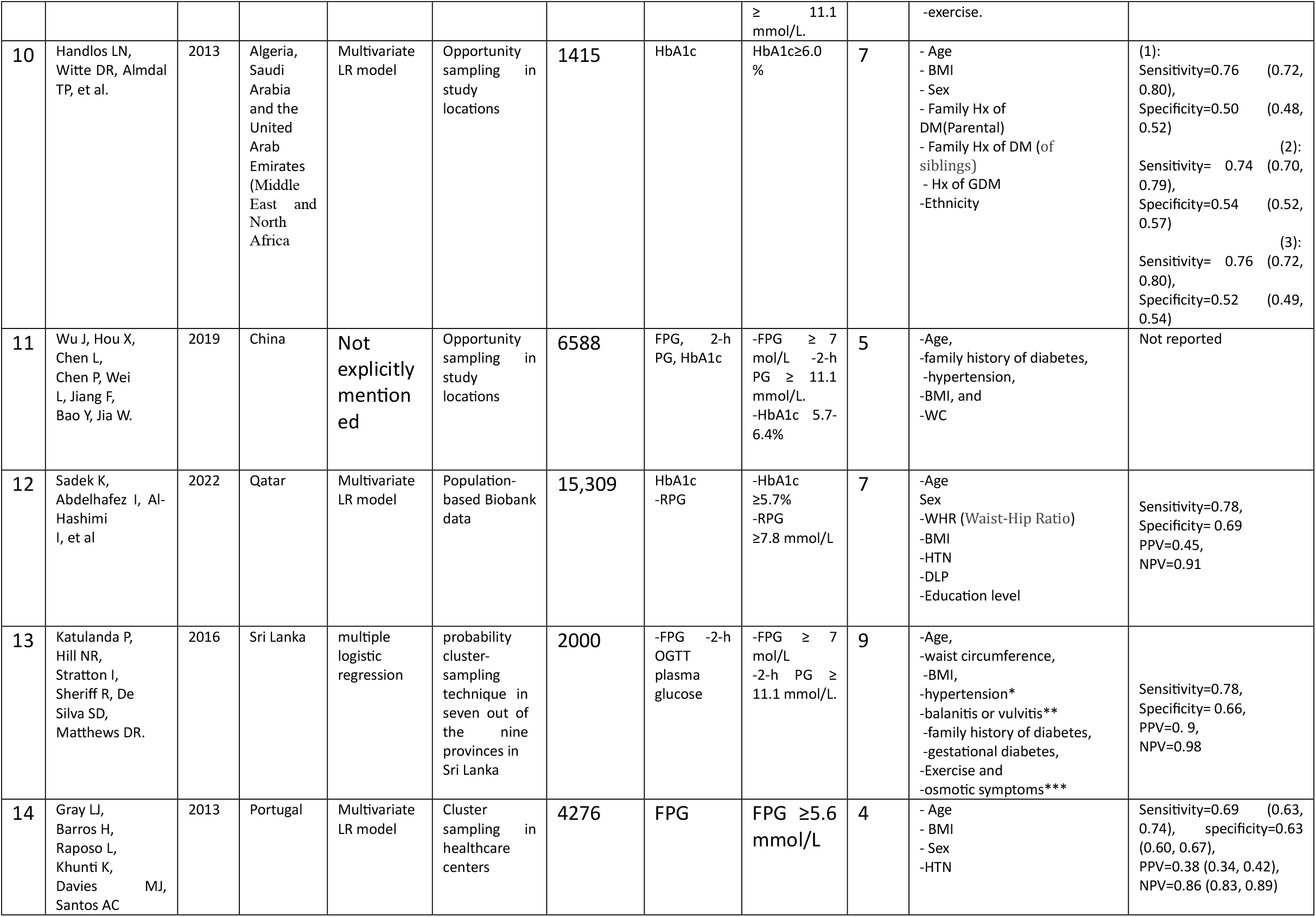
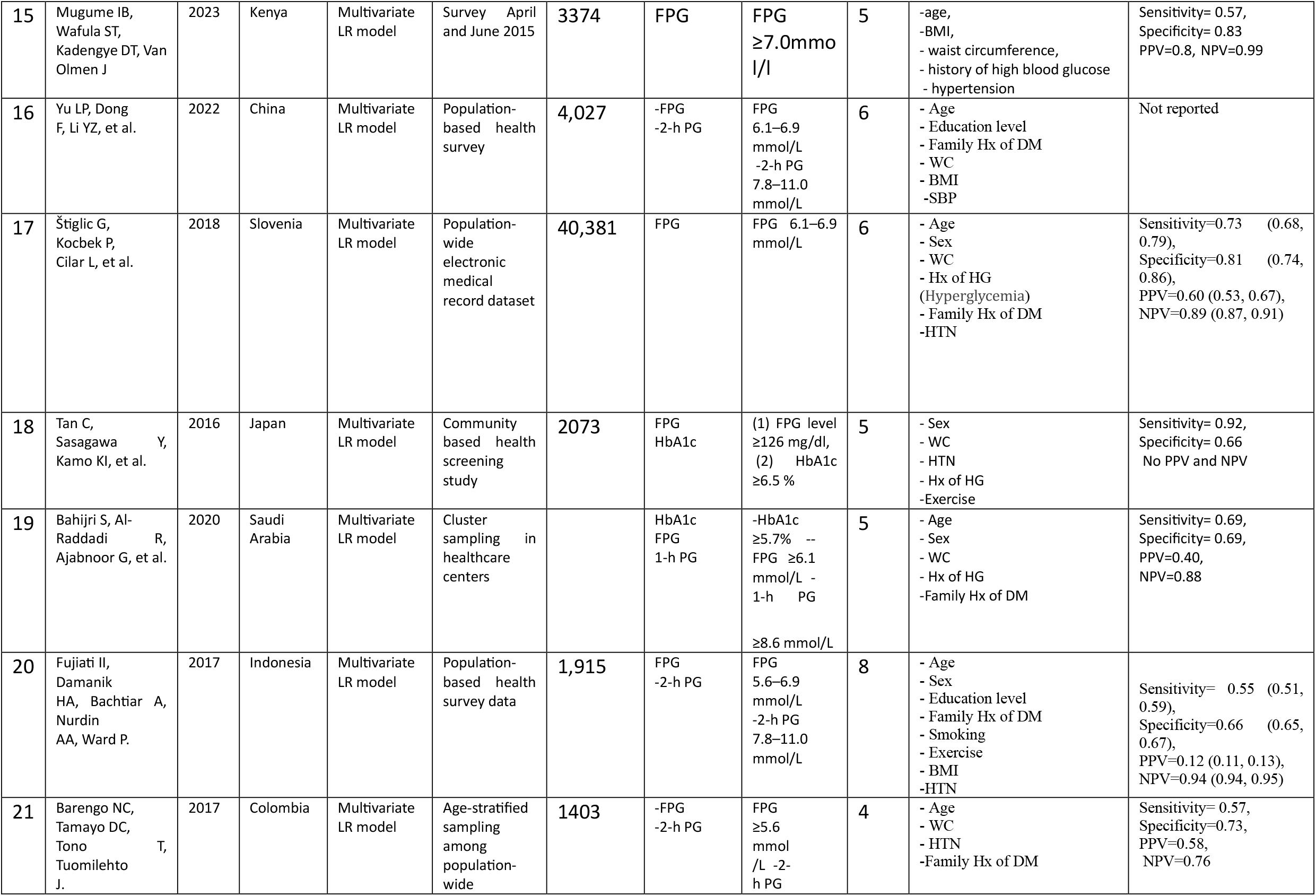
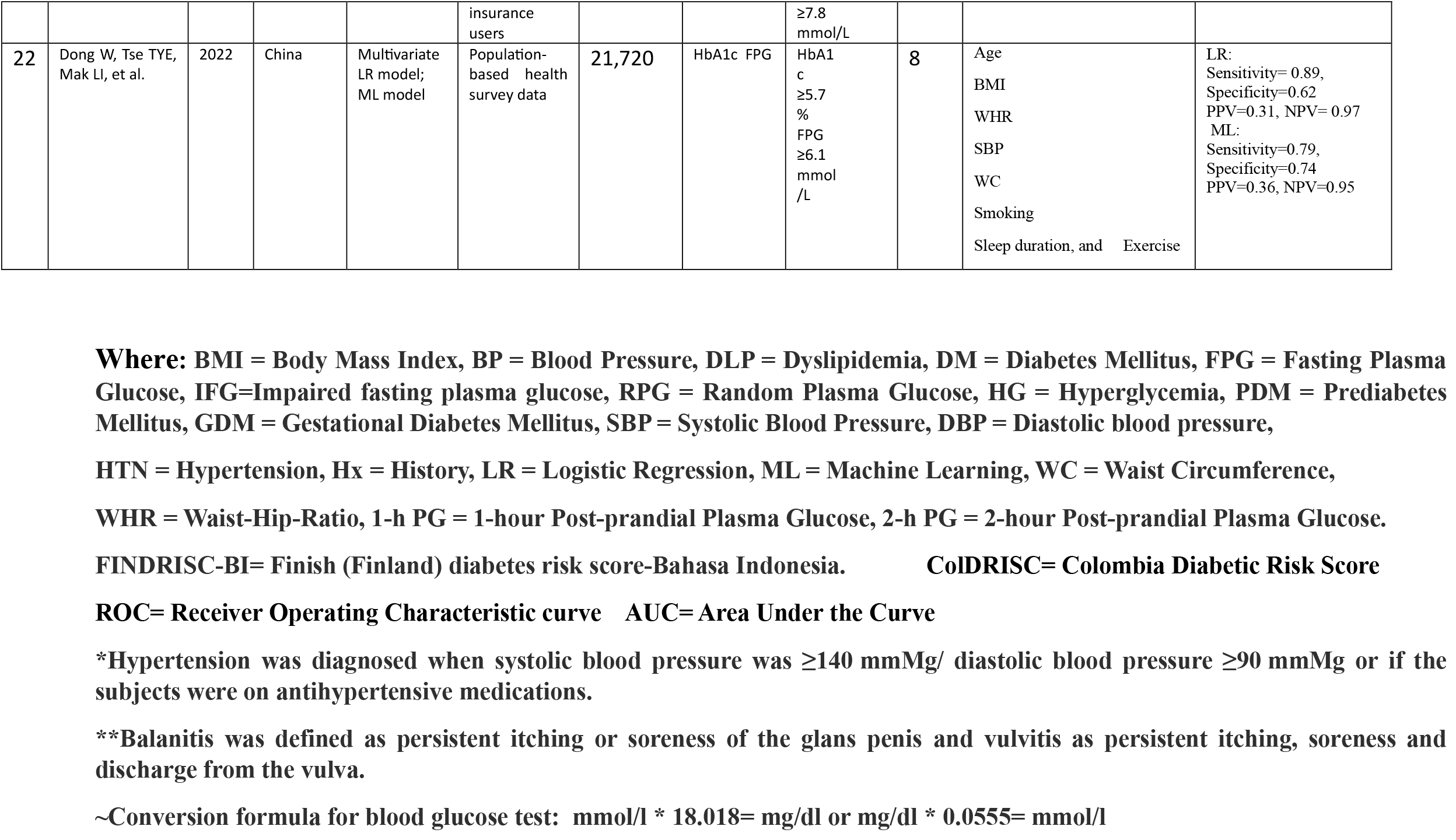
Summary of study variable characteristics and prediction tools of the included studies (n = 22)

| # | Author(s) | Year of publication | Origin/ country of origin | Methodology/methods | Sample source | sample size | The 'Gold standard' Outcome measure by | The 'Gold standard' reference of the Tool | # of Key findings | Key findings Predictors Tool | Predictive Power (Sen. (95% CI), Spe. (95% CI), PPV (95% CI), NPV (95% CI)) |
| --- | --- | --- | --- | --- | --- | --- | --- | --- | --- | --- | --- |
| 1 | Rokhman MR, Arifin B, Zulkarnain Z, et al | 2019 | Indonesia | Multivariate LR model | community members and government employees | 1,403 | FBG | -FBG <100mg/dL),<br>- prediabetes(pre-DM) (100-125mg/dL) and - T2DM (≥126mg/dL | 11 | -Age<br>-Gender<br>-Weight<br>-Height<br>-WC<br>-Exercise(>30m/d)<br>-Vegetable and fruit eating<br>-Hypertension*<br>-Hx of hyperglycemia<br>-Hx of diabetes in 1st degree Cuisine<br>-Hx of diabetes in 1st degree family | sensitivity=0.664; specificity = 0.67<br>No PPV and NPV |
| 2 | Fu Q, Sun M, Tang W, et al. | 2014 | China | Multivariate stepwise logistic regression | Community based health screening study | 7953 | 2-h PG | 2-h PG ≥7.8 mmol/L~ | 9 | - Age<br>- Height<br>- BMI<br>- WC<br>- Pulse<br>- SBP<br>- DLP<br>- HTN<br>----Family Hx of DM | Not reported |
| 3 | Memish ZA, Chang JL, Saeedi MY, Al Hamid MA, Abid O, Ali MK. | 2015 | Saudi Arabia | Multivariate LR model | Geographically stratified sampling in healthcare centers | 1435 | FPG/ 2-h PG | FPG ≥5.6 mmol/L<br>2-h PG ≥7.8 mmol/L | 4 | -Age<br>- Hx of GDM<br>- HTN<br>-WC | sensitivity=0.90, specificity= 0.84<br>No PPV and NPV |
| 4 | Rajput R, Garg K, Rajput M. | 2019 | India | Multivariate LR model | Opportunity sampling in study locations |  | FPG/ 2-h PG | FPG 5.6–6.9 mmol/L<br>2-h PG 7.8–11.0 mmol/L | 4 | - Age<br>- Family Hx of DM<br>- Waist-to-height ratio<br>-DBP | Sensitivity= 0.84 (0.78, 0.90),<br>Specificity=0.58(0.55, 0.62)<br>PPV=0.31 (0.27, 0.34),<br>NPV=0.94 (0.92, 0.96) |

|  |  |  |  |  |  |  |  |  |  |  |  |  |
| --- | --- | --- | --- | --- | --- | --- | --- | --- | --- | --- | --- | --- |
| 5 | Wang H, Liu T, Qiu Q, Ding P, He YH, Chen WQ. | 2015 | China | Multivariate LR model | Population-based health survey | 892 | FPG | FPG<br>6.1–6.9 mmol/L | 5,4 | **Men:**<br>- Age<br>- WC<br>- BMI<br>- Family Hx of DM<br>- HTN | **Women:**<br>- Age<br>- WC<br>- BMI<br>- Family Hx of DM | (1)<br>Men:<br>Sensitivity=0.73, specificity=0.64, PPV=0.13, NPV=0.97<br>Women:<br>Sensitivity=0.81, specificity=0.60, PPV=0.19, NPV=0.96<br>(2)<br>Men:<br>Sensitivity=0.79, specificity=0.51, PPV=0.06, NPV=0.99<br>Women:<br>Sensitivity=0.89, specificity=0.42, PPV=0.05, NPV=0.99<br>(3)<br>Men:<br>Sensitivity=0.31, specificity=0.49, PPV=0.02, NPV=0.96<br>Women:<br>Sensitivity=0.42, specificity=0.59, PPV=0.04, NPV=0.96 |
| 6 | Henjum S, Hjellset VT, Andersen E, Flaaten MØ, Morseth MS. | 2022 | Algeria Saharawi refugees in Algeria | Multivariate LR model | Opportunity sampling in study locations | 6197 | HbA1c | HbA1c ≥5.7% | 3 | - Age<br>- BMI<br>- WC |  | Sensitivity=0.89, Specificity= 0.65, PPV=0.28, NPV=0.97 |
| 7 | Zhang, Lin, L., Xu, X., Wu, X., Jin, Q., & Liu, H. | 2019 | China | multivariable logistic regression model | Chinese communities | 308 | FPG HbA1c | -FPG ≥ 126 mg/dL (≥ 7.0 mmol/L) or - glycated hemoglobin (HbA1c) ≥ 6.5% | 7 | - age,<br>-family history of diabetes,<br>-hypertension,<br>-waist circumference,<br>-body mass index,<br>-smoking,<br>-daily consumption of vegetables, and fruits. |  | Sensitivity=0.83, Specificity= 0.60<br>No PPV and NPV |
| 8 | Abbas M, Mall R, Errafii K, et al. | 2021 | Qatar | Multivariate LR model ML | Population-based Biobank data | 1432 | HbA1c | HbA1c 5.7-6.4% | 5 | - Age<br>- BMI<br>- HTN<br>- Sex<br>- WC |  | Sensitivity=0.86 (0.83, 0.89), Specificity=0.58 (0.55, 0.61), PPV=0.50 (0.46, 0.53), NPV=0.90 (0.87, 0.92) |
| 9 | Woo YC, Lee CH, Fong CHY, Tso AWK, Cheung BMY, Lam KSL. | 2017 | China | LR model | Invited randomly by their telephone numbers | 5814 | FBG 2-h PG | fasting glucose ≥ 7 mmol/L or 2hour post oral glucose tolerance test glucose | 8 | -age,<br>-waist circumference,<br>-gestational diabetes,<br>-height,<br>-race/ethnicity,<br>-hypertension,<br>-family history of Diabetes, |  | Sensitivity=0.81, specificity=0.41<br>PPV=0.09, NPV=0.97 |
|  |  |  |  |  |  |  |  | ≥ 11.1 mmol/L. |  | -exercise. |  |
| 10 | Handlos LN, Witte DR, Almdal TP, et al. | 2013 | Algeria, Saudi Arabia and the United Arab Emirates (Middle East and North Africa | Multivariate LR model | Opportunity sampling in study locations | 1415 | HbA1c | HbA1c≥6.0 % | 7 | - Age<br>- BMI<br>- Sex<br>- Family Hx of DM(Parental)<br>- Family Hx of DM (of siblings)<br>- Hx of GDM<br>- Ethnicity | (1):<br>Sensitivity=0.76 (0.72, 0.80),<br>Specificity=0.50 (0.48, 0.52)<br>(2):<br>Sensitivity= 0.74 (0.70, 0.79),<br>Specificity=0.54 (0.52, 0.57)<br>(3):<br>Sensitivity= 0.76 (0.72, 0.80),<br>Specificity=0.52 (0.49, 0.54) |
| 11 | Wu J, Hou X, Chen L, Chen P, Wei L, Jiang F, Bao Y, Jia W. | 2019 | China | Not explicitly mentioned | Opportunity sampling in study locations | 6588 | FPG, 2-h PG, HbA1c | -FPG ≥ 7 mol/L<br>-2-h PG ≥ 11.1 mmol/L.<br>-HbA1c 5.7-6.4% | 5 | -Age,<br>-family history of diabetes,<br>-hypertension,<br>-BMI, and<br>-WC | Not reported |
| 12 | Sadek K, Abdelhafez I, Al-Hashimi I, et al | 2022 | Qatar | Multivariate LR model | Population-based Biobank data | 15,309 | HbA1c<br>-RPG | -HbA1c ≥5.7%<br>-RPG ≥7.8 mmol/L | 7 | -Age<br>Sex<br>-WHR (Waist-Hip Ratio)<br>-BMI<br>-HTN<br>-DLP<br>-Education level | Sensitivity=0.78,<br>Specificity= 0.69<br>PPV=0.45,<br>NPV=0.91 |
| 13 | Katulanda P, Hill NR, Stratton I, Sheriff R, De Silva SD, Matthews DR. | 2016 | Sri Lanka | multiple logistic regression | probability cluster-sampling technique in seven out of the nine provinces in Sri Lanka | 2000 | -FPG -2-h OGTT plasma glucose | -FPG ≥ 7 mol/L<br>-2-h PG ≥ 11.1 mmol/L. | 9 | -Age,<br>-waist circumference,<br>-BMI,<br>-hypertension*<br>-balanitis or vulvitis**<br>-family history of diabetes,<br>-gestational diabetes,<br>-Exercise and<br>-osmotic symptoms*** | Sensitivity=0.78,<br>Specificity= 0.66,<br>PPV=0. 9,<br>NPV=0.98 |
| 14 | Gray LJ, Barros H, Raposo L, Khunti K, Davies MJ, Santos AC | 2013 | Portugal | Multivariate LR model | Cluster sampling in healthcare centers | 4276 | FPG | FPG ≥5.6 mmol/L | 4 | - Age<br>- BMI<br>- Sex<br>-HTN | Sensitivity=0.69 (0.63, 0.74), specificity=0.63 (0.60, 0.67),<br>PPV=0.38 (0.34, 0.42),<br>NPV=0.86 (0.83, 0.89) |
| 15 | Mugume IB, Wafula ST, Kadengye DT, Van Olmen J | 2023 | Kenya | Multivariate LR model | Survey April and June 2015 | 3374 | FPG | FPG $\geq 7.0$ mmol/L | 5 | -age, -BMI, - waist circumference, - history of high blood glucose - hypertension | Sensitivity= 0.57, Specificity= 0.83 PPV=0.8, NPV=0.99 |
| 16 | Yu LP, Dong F, Li YZ, et al. | 2022 | China | Multivariate LR model | Population-based health survey | 4,027 | -FPG -2-h PG | FPG 6.1–6.9 mmol/L -2-h PG 7.8–11.0 mmol/L | 6 | - Age - Education level - Family Hx of DM - WC - BMI -SBP | Not reported |
| 17 | Štiglic G, Kocbek P, Cilar L, et al. | 2018 | Slovenia | Multivariate LR model | Population-wide electronic medical record dataset | 40,381 | FPG | FPG 6.1–6.9 mmol/L | 6 | - Age - Sex - WC - Hx of HG (Hyperglycemia) - Family Hx of DM -HTN | Sensitivity=0.73 (0.68, 0.79), Specificity=0.81 (0.74, 0.86), PPV=0.60 (0.53, 0.67), NPV=0.89 (0.87, 0.91) |
| 18 | Tan C, Sasagawa Y, Kamo KI, et al. | 2016 | Japan | Multivariate LR model | Community based health screening study | 2073 | FPG HbA1c | (1) FPG level $\geq 126$ mg/dl, (2) HbA1c $\geq 6.5$ % | 5 | - Sex - WC - HTN - Hx of HG -Exercise | Sensitivity= 0.92, Specificity= 0.66 No PPV and NPV |
| 19 | Bahijri S, Al-Raddadi R, Ajabnoor G, et al. | 2020 | Saudi Arabia | Multivariate LR model | Cluster sampling in healthcare centers | | HbA1c FPG 1-h PG | -HbA1c $\geq 5.7$ % -- FPG $\geq 6.1$ mmol/L - 1-h PG $\geq 8.6$ mmol/L | 5 | - Age - Sex - WC - Hx of HG -Family Hx of DM | Sensitivity= 0.69, Specificity= 0.69, PPV=0.40, NPV=0.88 |
| 20 | Fujiati II, Damanik HA, Bachtiar A, Nurdin AA, Ward P. | 2017 | Indonesia | Multivariate LR model | Population-based health survey data | 1,915 | FPG -2-h PG | FPG 5.6–6.9 mmol/L -2-h PG 7.8–11.0 mmol/L | 8 | - Age - Sex - Education level - Family Hx of DM - Smoking - Exercise - BMI -HTN | Sensitivity= 0.55 (0.51, 0.59), Specificity=0.66 (0.65, 0.67), PPV=0.12 (0.11, 0.13), NPV=0.94 (0.94, 0.95) |
| 21 | Barengo NC, Tamayo DC, Tono T, Tuomilehto J. | 2017 | Colombia | Multivariate LR model | Age-stratified sampling among population-wide | 1403 | -FPG -2-h PG | FPG $\geq 5.6$ mmol/L -2-h PG | 4 | - Age - WC - HTN -Family Hx of DM | Sensitivity= 0.57, Specificity=0.73, PPV=0.58, NPV=0.76 |
|  |  |  |  |  | insurance users |  |  | ≥7.8 mmol/L |  |  |  |
| 22 | Dong W, Tse TYE, Mak LI, et al. | 2022 | China | Multivariate LR model; ML model | Population-based health survey data | 21,720 | HbA1c FPG | HbA1c ≥5.7 %<br>FPG ≥6.1 mmol/L | 8 | Age<br>BMI<br>WHR<br>SBP<br>WC<br>Smoking<br>Sleep duration, and Exercise | LR:<br>Sensitivity= 0.89,<br>Specificity=0.62<br>PPV=0.31, NPV= 0.97<br>ML:<br>Sensitivity=0.79,<br>Specificity=0.74<br>PPV=0.36, NPV=0.95 |

**Where:** BMI = Body Mass Index, BP = Blood Pressure, DLP = Dyslipidemia, DM = Diabetes Mellitus, FPG = Fasting Plasma Glucose, IFG=Impaired fasting plasma glucose, RPG = Random Plasma Glucose, HG = Hyperglycemia, PDM = Prediabetes Mellitus, GDM = Gestational Diabetes Mellitus, SBP = Systolic Blood Pressure, DBP = Diastolic blood pressure,
HTN = Hypertension, Hx = History, LR = Logistic Regression, ML = Machine Learning, WC = Waist Circumference,
WHR = Waist-Hip-Ratio, 1-h PG = 1-hour Post-prandial Plasma Glucose, 2-h PG = 2-hour Post-prandial Plasma Glucose.
FINDRISC-BI= Finish (Finland) diabetes risk score-Bahasa Indonesia.
ColDRISC= Colombia Diabetic Risk Score
ROC= Receiver Operating Characteristic curve AUC= Area Under the Curve
\*Hypertension was diagnosed when systolic blood pressure was $\geq 140$ mmHg/ diastolic blood pressure $\geq 90$ mmHg or if the subjects were on antihypertensive medications.
\*\*Balanitis was defined as persistent itching or soreness of the glans penis and vulvitis as persistent itching, soreness and discharge from the vulva.
~Conversion formula for blood glucose test: mmol/l \* 18.018= mg/dl or mg/dl \* 0.0555= mmol/l

This review included studies conducted from 2013 to 2023, 36% studies being conducted in 2019 and 2022. Generally, 50% of the article published in the last 5 years. (see fig 2)

**Figure 2.**
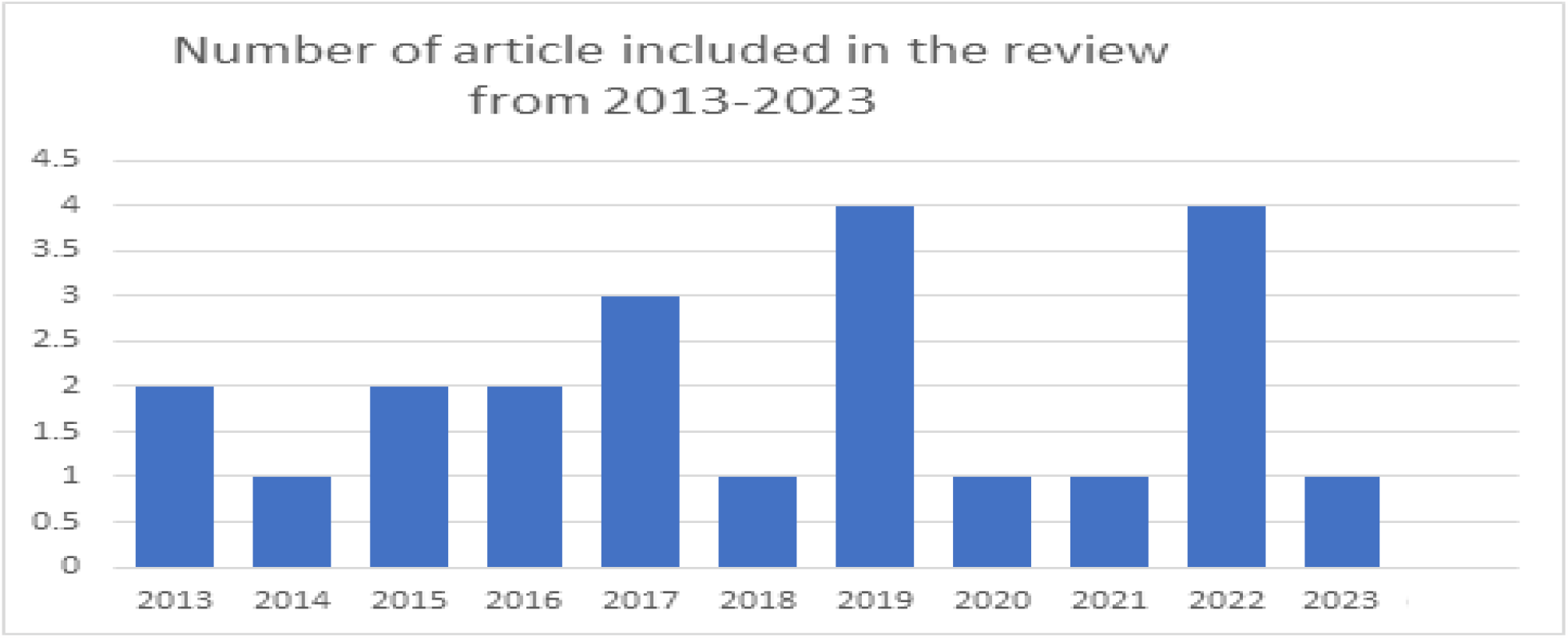
The number of the included studies in each year from 2013 to 2023.

The included articles are from 13 countries, and the majority were conducted in China (32%) ^4,7,9,11,13,18,25^ followed by three articles from Saudi Arabia (13.6%),^5,12,21^ two articles from Indonesia, Algeria and Qatar (9%).^3,6,10,14,22^ Single paper from the rest of 8 countries (each 5%). (See fig. 3)

**Fig 3:**
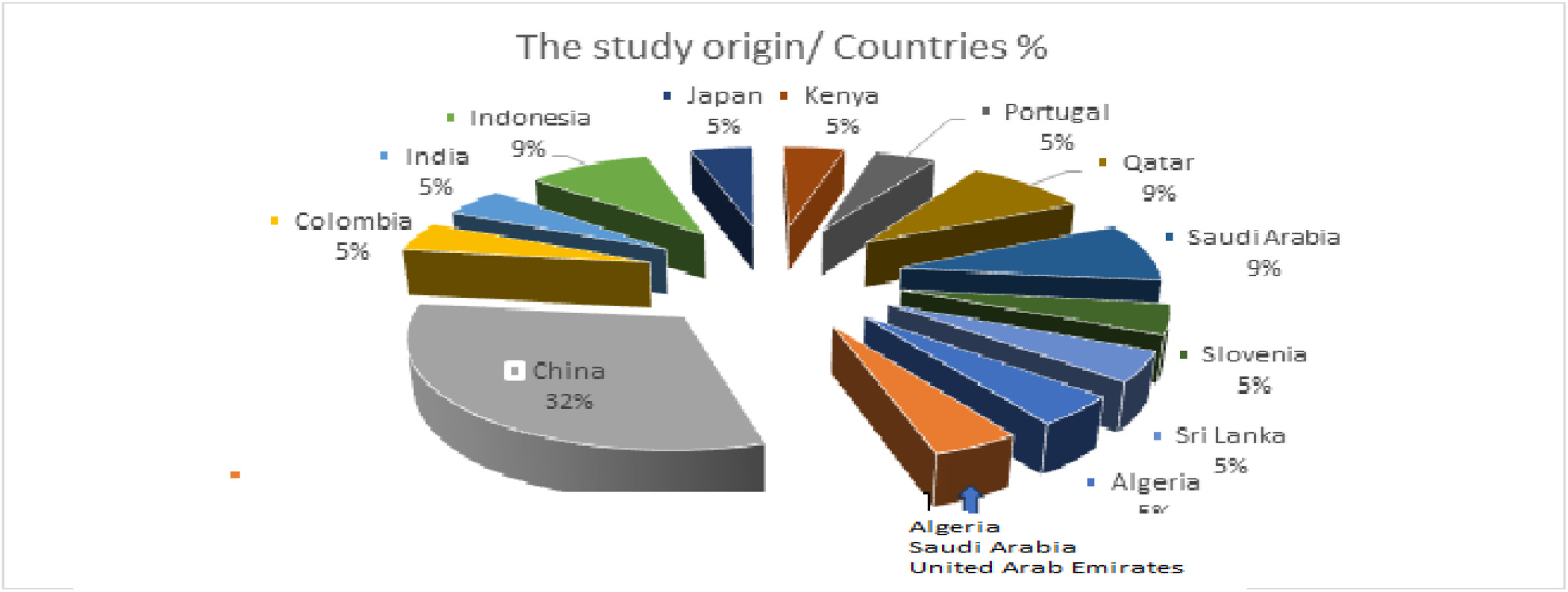
shows Countries where the included studies were conducted.

The authors of the articles got data from four sources. Eleven studies’ data source is from community survey (50%),^4,5,6,8,9,12,16,17,20,21,23^ Eight studies used data from databank (36.4%), ^3,7,10,14,18,19,22,24^ another two studies collected data by random sampling (9%)^11,15^ and the rest one study collected data by non-random sampling (Convenience sampling) (4.5%)^13^to develop their risk prediction tools. The median sample size was 1,403 and ranged from 308 to 40,381. (See table 1)

The key risk Predictors of pre-Diabetes utilization frequencies to develop the Tools across the included articles are summarized in Figure 4. A total of 24 key risk predictors were used to develop different types of tools. Among the key risk predictors repeatedly used by the Tool developer are listed, and the median is three, ranging from 1 to 20 (see Figure 4). Age was the most common predictor to predict pre-DM as it was included in 20 studies (90.9%). Other commonly included factors were Hypertension and waist circumference in 16 studies (72.7%), BMI in 15 studies (68.2%), family history of DM in 14 studies (63.6%), Gender (sex) in 8 studies (36.4%), Exercise in 6 studies (27%). The rest of the parameters were used in 1 to 5 studies. (see fig. 4)

**Figure 4.**
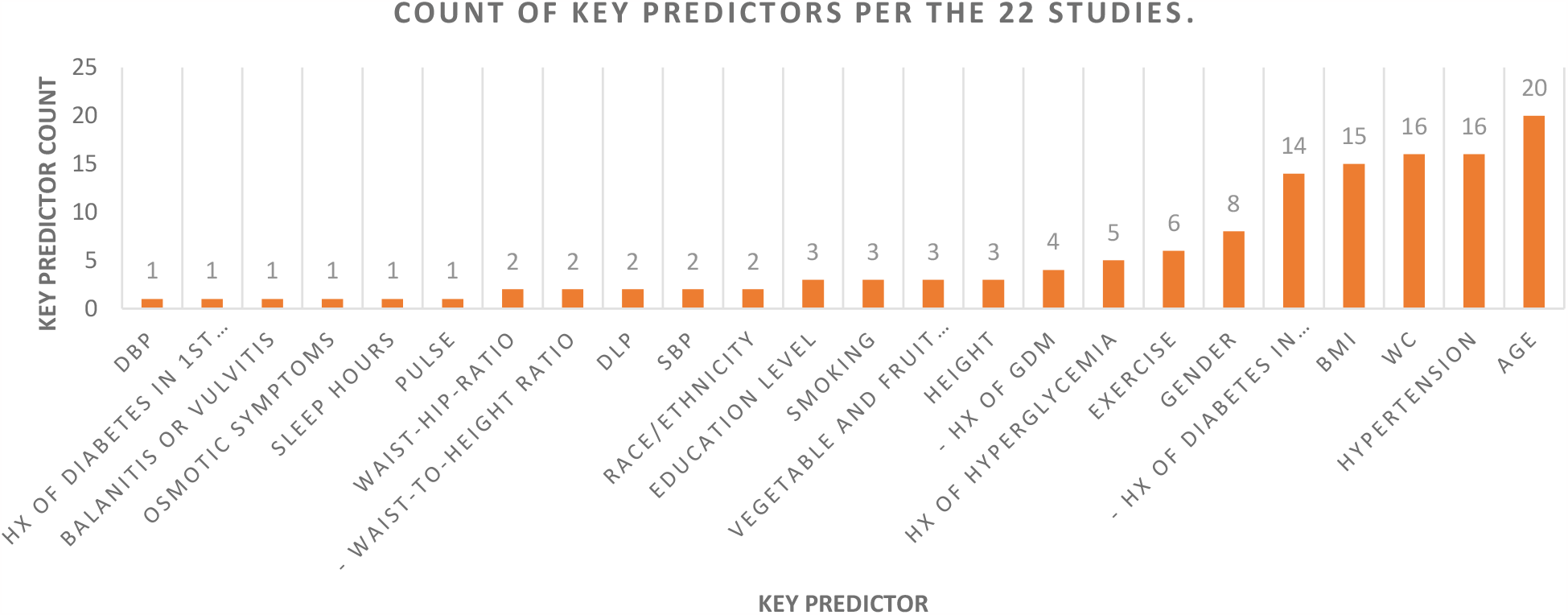
The number of studies utilized each key risk predictor to develop the Tools

Among the key predictors, less commonly used include pulse,^4^ sleep duration,^24^osmotic symptoms,^15^ Balanitis or vulvitis,^15^ and DBP.^6^

The methodology used to prepare the Tools was adequately reported in most of the studies. The researchers used Logistic regression (LR) Methods for Tool development in 20 studies (90%). Two studies used machine learning (ML) in addition to LR.^10,24^ These two articles reported no significant difference in predictive performances in the tools developed by LR and ML methods.^10,24^

The performance of each tool evaluated by ROC curve and the value of the AUC ranges from 0.65-0.93 by. The lowest AUC by a tool [Age, Height, BMI, WC, Pulse, SBP, DLP, HTN, Family Hx of DM] developed by Fu Q, Sun M, Tang W, et al. and the highest is [Sex, WC, HTN, Hx of HG, Exercise] developed by Tan C, Sasagawa Y, Kamo KI, et al. (see table 2 and Figure 5 and 6)

**Table 2:**
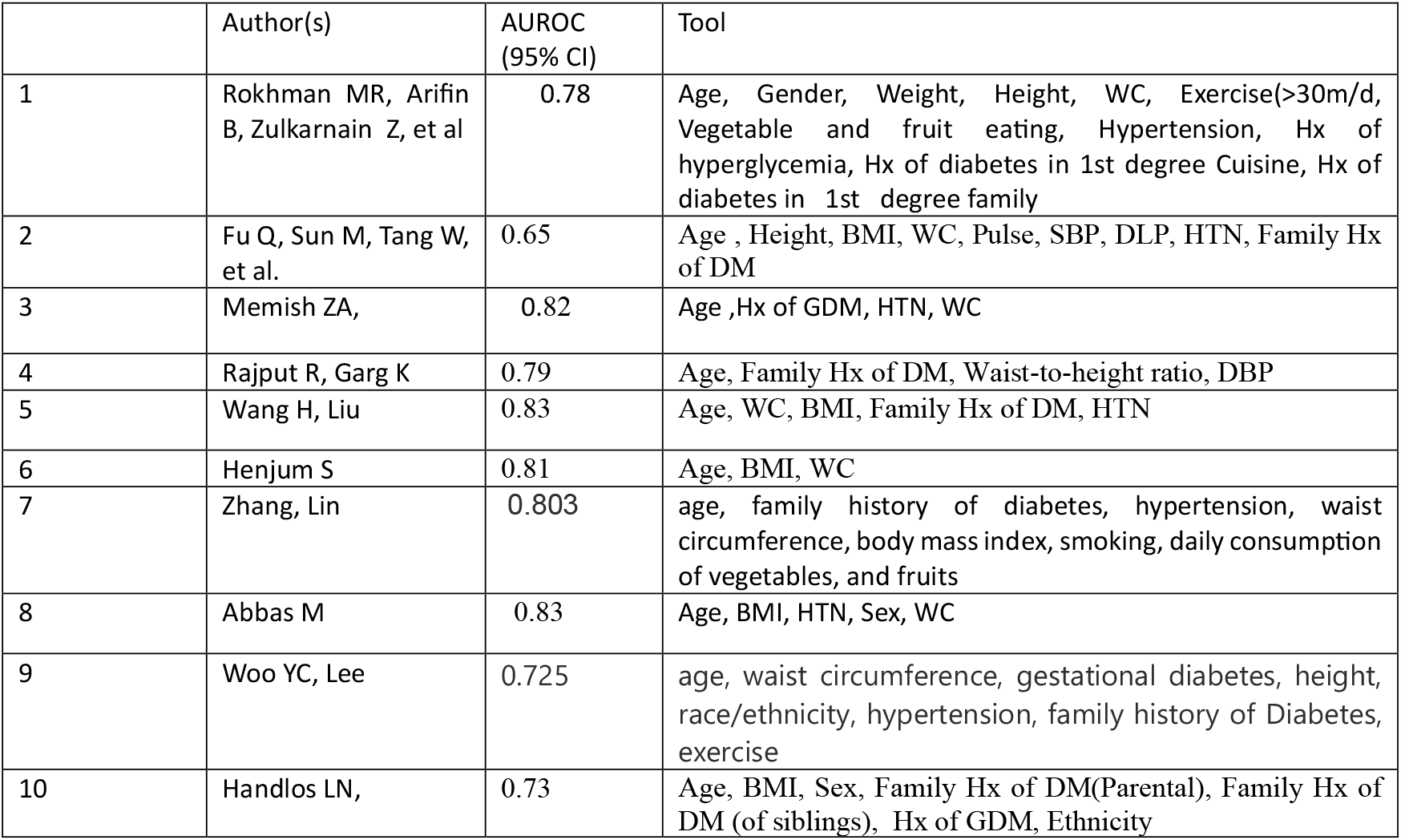
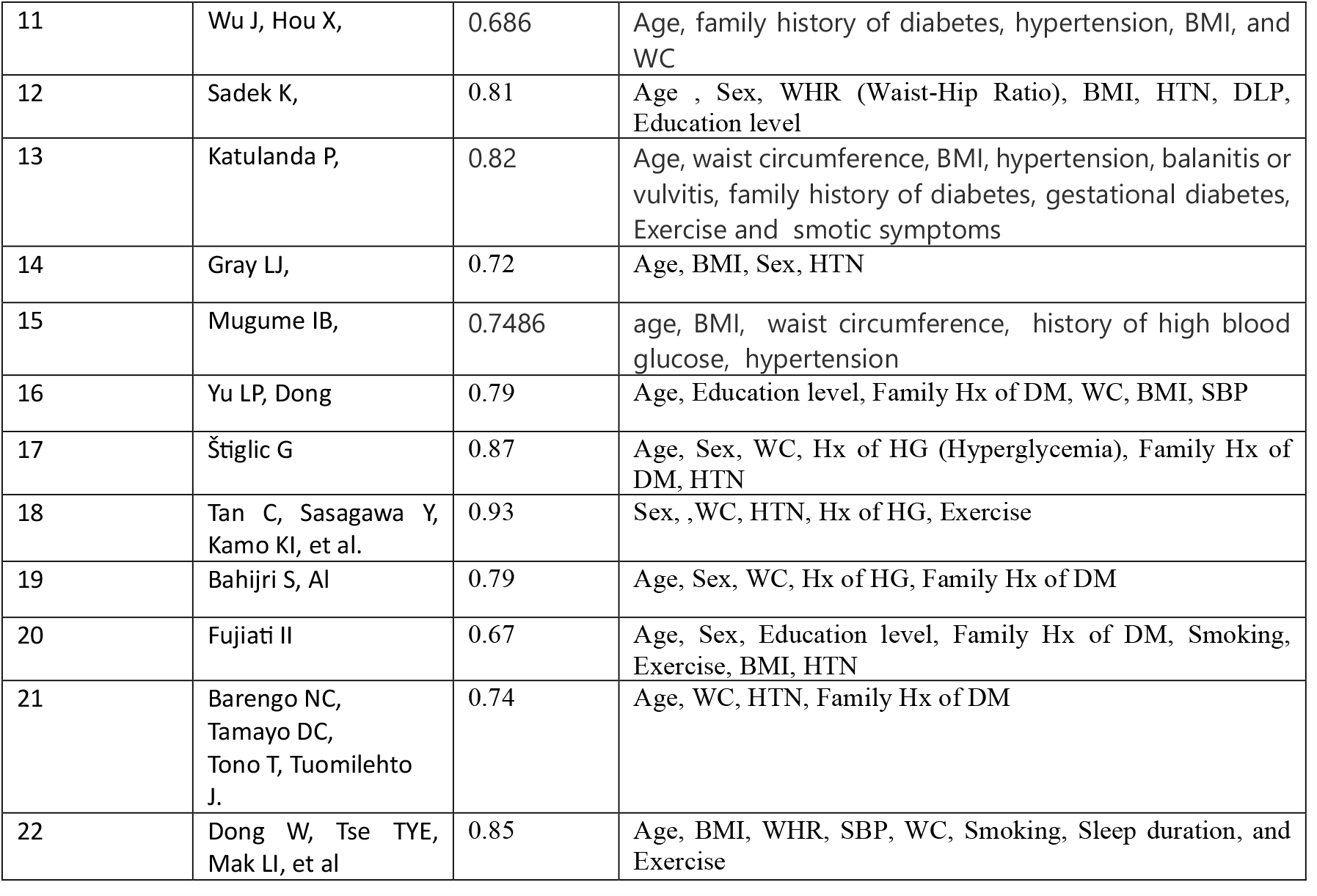
Table shows the result of AUROC / performance evaluation/ of the 22 Tools.

**Figure 5.**
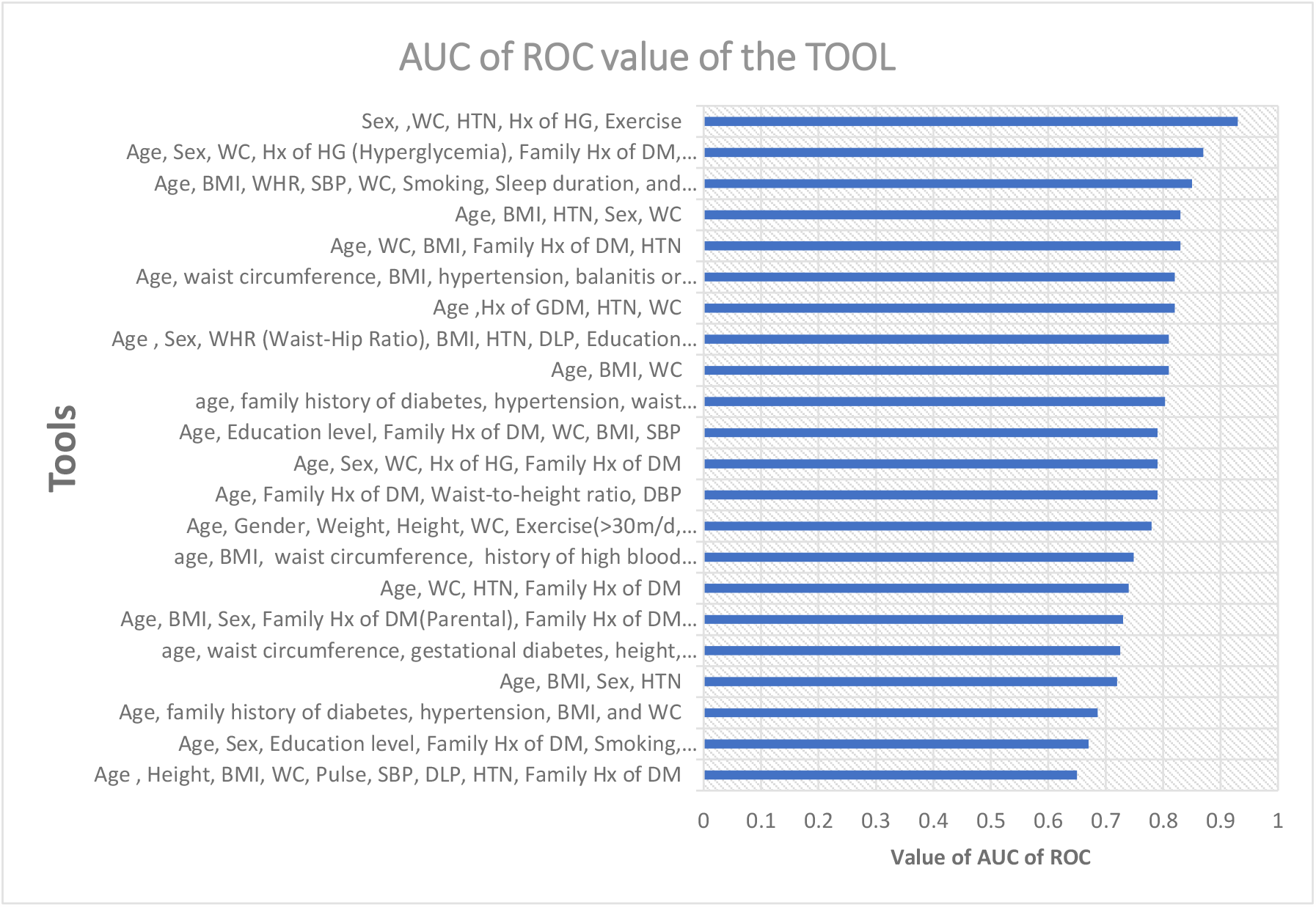
shows the AUC of ROC value of the TOOL

**Figure 6.**
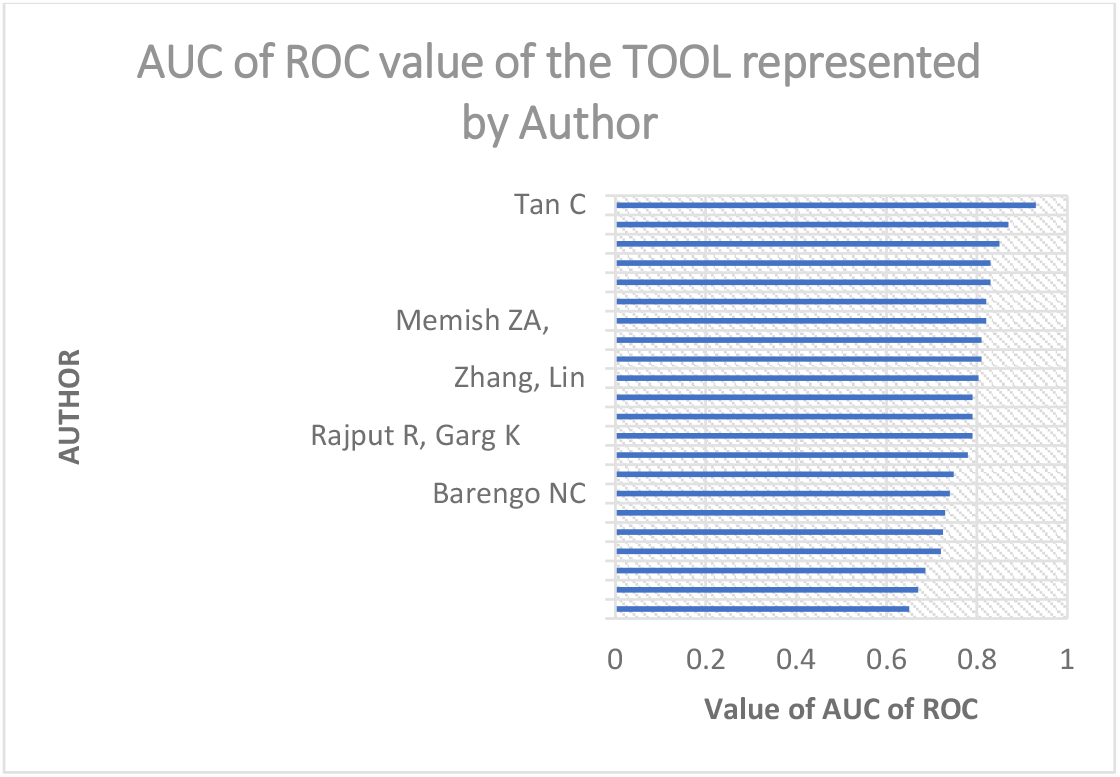
shows the AUC of ROC value of the TOOL represented by Authors

The authors of the articles described the predictive power of the screening tool using Sensitivity, Specificity, PPV and NPV metrics. Sensitivity and specificity are concerned with the accuracy of the screening tool in reference to the ‘gold standard’ value. Sensitivities and specificities of the tools were performed either during development or at the time of validation. The value of Sensitivity and Specificity of the developed screening Tools are displayed in table 1 and Figures 7,8 and 9.

**Figure 7.**
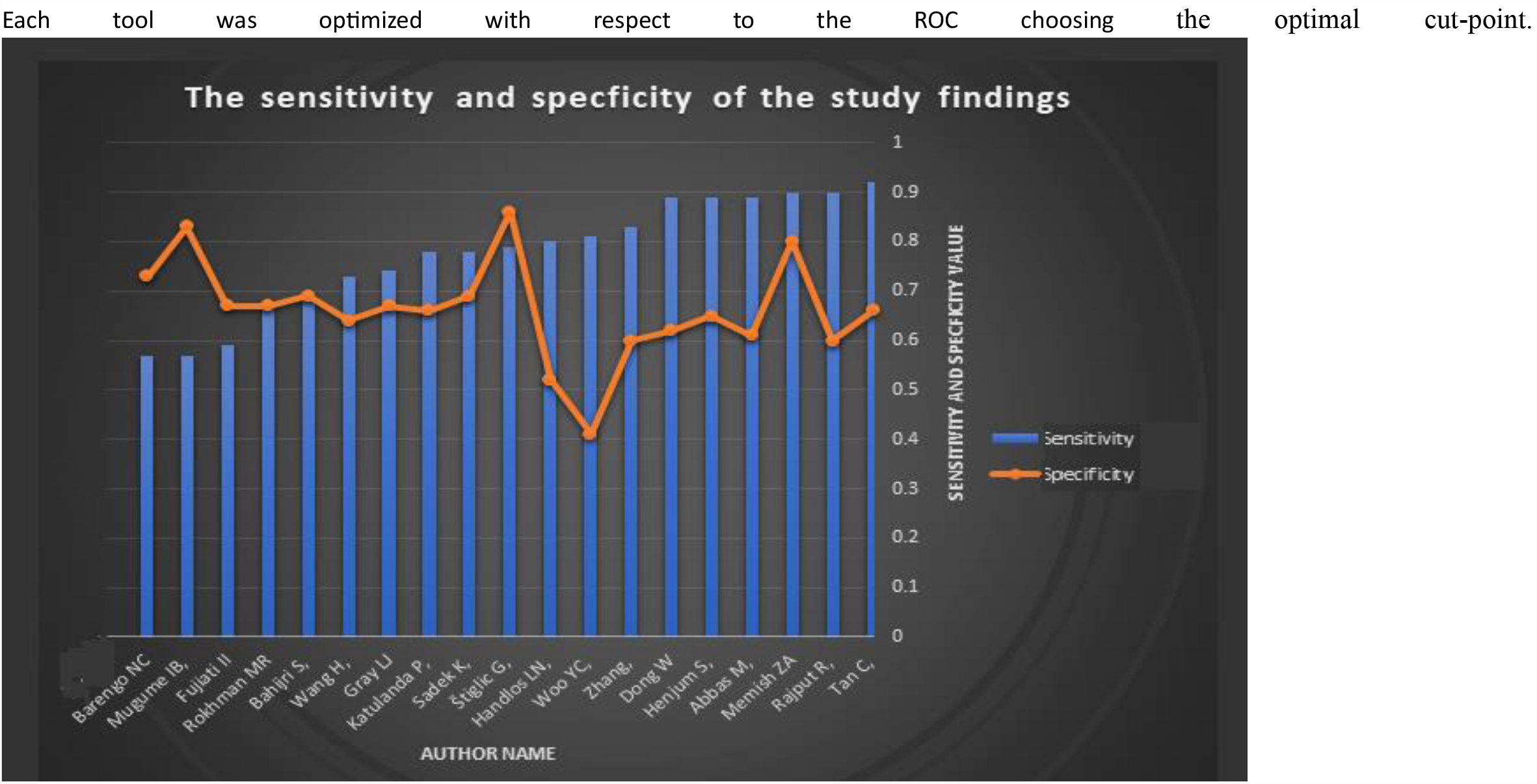
Sensitivity and Specificity of Predictive Tool represented by the Authors

**Figure 8.**
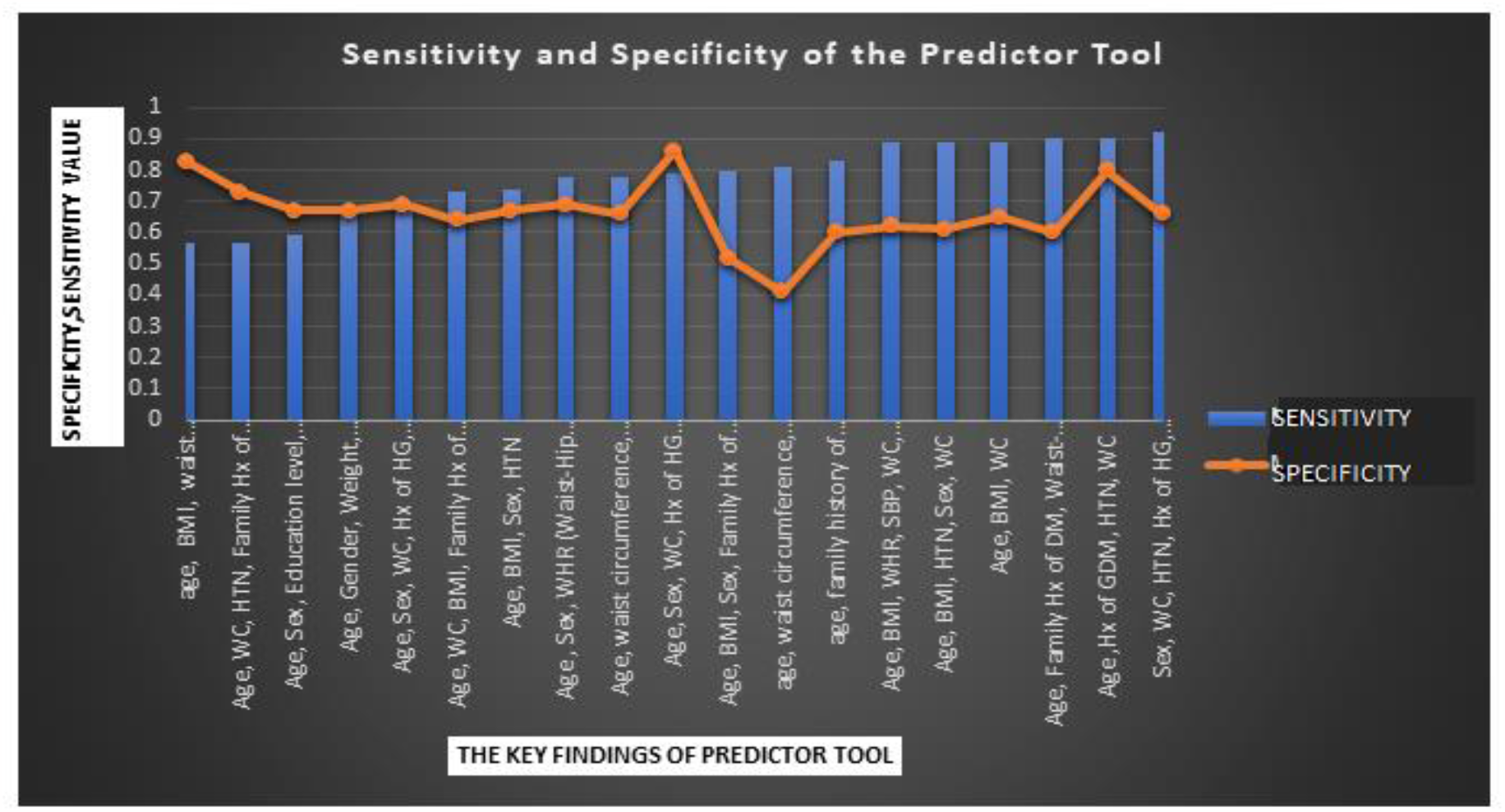
Sensitivity and Specificity of Key Finding representing the Predictor Tool

**Figure 9.**
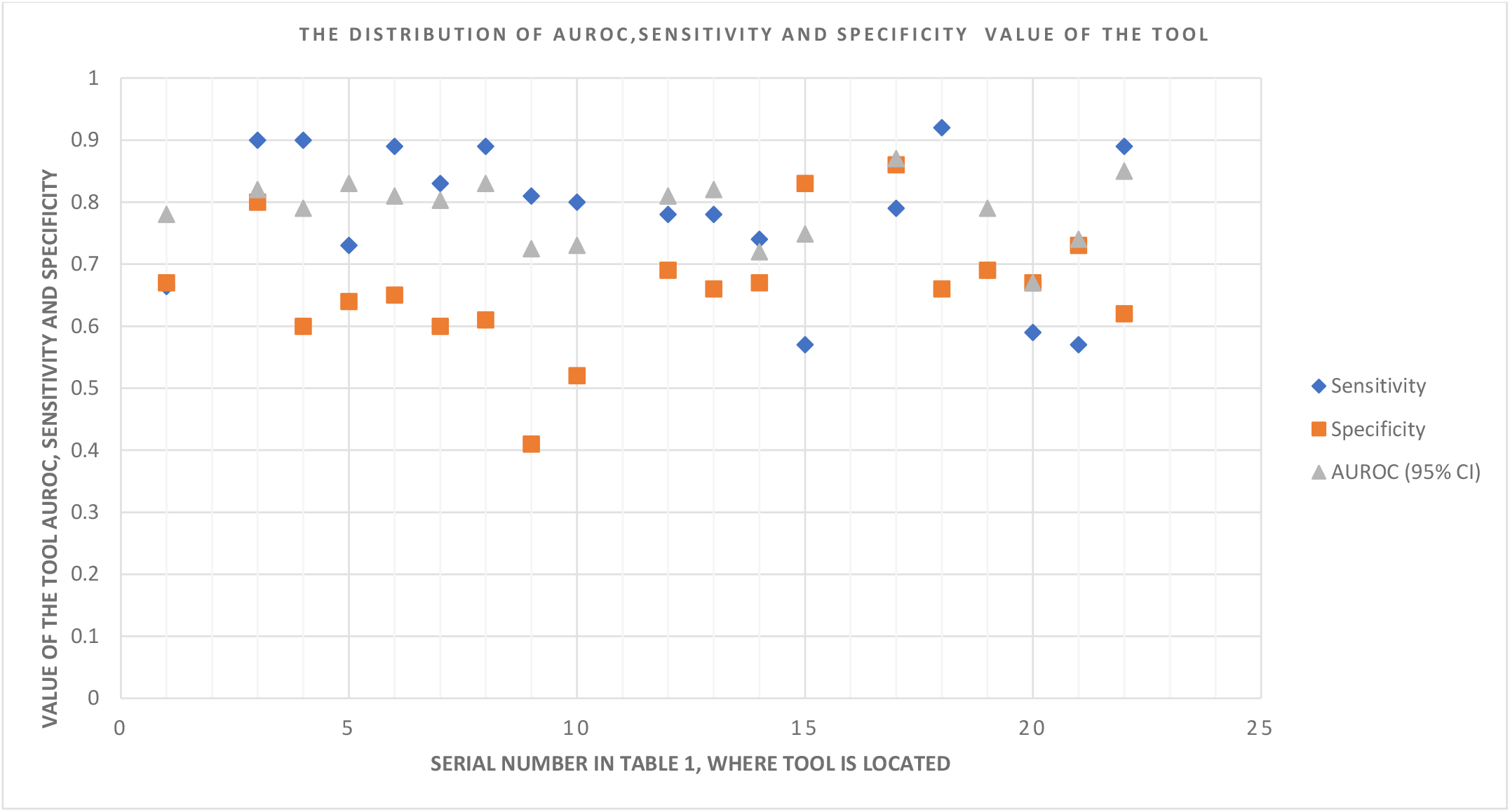
SCATTER PLOT OF AUROC, SENSITIVITY, SPECIFICITY VALUE AND THEIR CORRELATION (OF THE TOOL)

Fourteen studies (63.6%)^6,7,8,10,11,14,15,16,17,19,21,22,23,24^ reported the performance of their prediction power using sensitivity, specificity, PPV, and NPV. However, in five studies (22.7%), ^3,5,9,12,20^ PPV and NPV were not reported, and the remaining three studies (13.6%) did not report all.^4,13,18^

To evaluate the predictive power of the developed noninvasive screening tool, the biochemical parameters utilized as the ‘Gold standard’ varied a lot among the studies. For instance, eight studies (36.4%) used FBS and 2hr PG,^5,6,11,15,18,20,22,23^ five studies (22.7%) used only FBS,^3,7,16,17,19^ two studies used FBS and HbA1c (9%),^9,24^ three studies (13.6%) used only HbA1C,^8,10,12^ one study (4.5%) used FBS, 1h PG and HbA1C,^21^ one study (4.5%) used FBS, 2h PG and HbA1c,^13^ one study (4.5%) used RBS and HbA1C,^14^ and one study (4.5%) used 2h PG to define pre-DM and DM.^4^ to calibrate the developed Tools. (See Table 1)

Though the evaluation method differs from study to study, generally, the author used the ‘gold standard’ references to ensure the Tool’s accuracy. Among the included studies’ predictive Tools, the highest sensitivity finding is 0.92 (92%) and the lowest is 0.57 (57%). Sex, WC, HTN, Hx of HG, and Exercise represent the highest sensitivity finding tool. This Tool was developed by Tan C, Sasagawa Y, Kamo KI, et al.^20^ On the other hand, the lowest sensitivity result is recorded on the Tool’s key risk predictive parameters, including ‘Age, WC, HTN, Family Hx of DM’ developed by Barengo NC, Tamayo DC, Tono T, Tuomilehto J.^23^

On the contrary, the studies’ predictive Tools Specificity ranges from 0.83 (83%) to 0.41 (41%), highest and lowest, respectively. The highest specificity is recorded by ‘age, BMI, waist circumference, history of high blood glucose, history of hypertension’ tool developed by Mugume IB, Wafula ST, Kadengye DT, Van Olmen J.^17^ The lowest Specificity test result recorded by the tool’ age, waist circumference, gestational diabetes, height, race/ethnicity, hypertension, family history of Diabetes, exercise’ developed by Woo YC, Lee CH, Fong CHY, Tso AWK, Cheung BMY, and Lam KSL^.11^ (See table 1, and Figure 7,8 and 9)

The developed Tools’ positive predictive value (PPV) and negative predictive value (NPV) indicate the Tool’s effectiveness in categorizing people as having or not having a Diabetes metabolic abnormality.^25,26^ The developed Tool has PPV ranging from 0.09 (9%) to 0.80 (80%). The highest PPV having Tool is ‘age, BMI, waist circumference, history of high blood glucose, history of hypertension’ developed by Mugume IB, Wafula ST, Kadengye DT, Van Olmen J.^17^ On the other hand, the lowest PPV tool is ‘age, waist circumference, gestational diabetes, height, race/ethnicity, hypertension, family history of Diabetes, and exercise’ developed by Woo YC, Lee CH, Fong CHY, Tso AWK, Cheung BMY, and Lam KSL.^11^

The NPV of the developed Tool ranges from 0.76 (76%) to 0.99 (99%). The highest NPV having Tool is ‘age, BMI, waist circumference, history of high blood glucose, and history of hypertension’ developed by Mugume IB, Wafula ST, Kadengye DT, Van Olmen J.^17^ On the other hand, the lowest NPV is 0.76 (76%). The Tool is ‘Age, WC, HTN, Family Hx of DM’ developed by Barengo NC, Tamayo DC, Tono T, Tuomilehto J.^23^ (See table 1, and figures 10 and 11)

**Figure 10.**
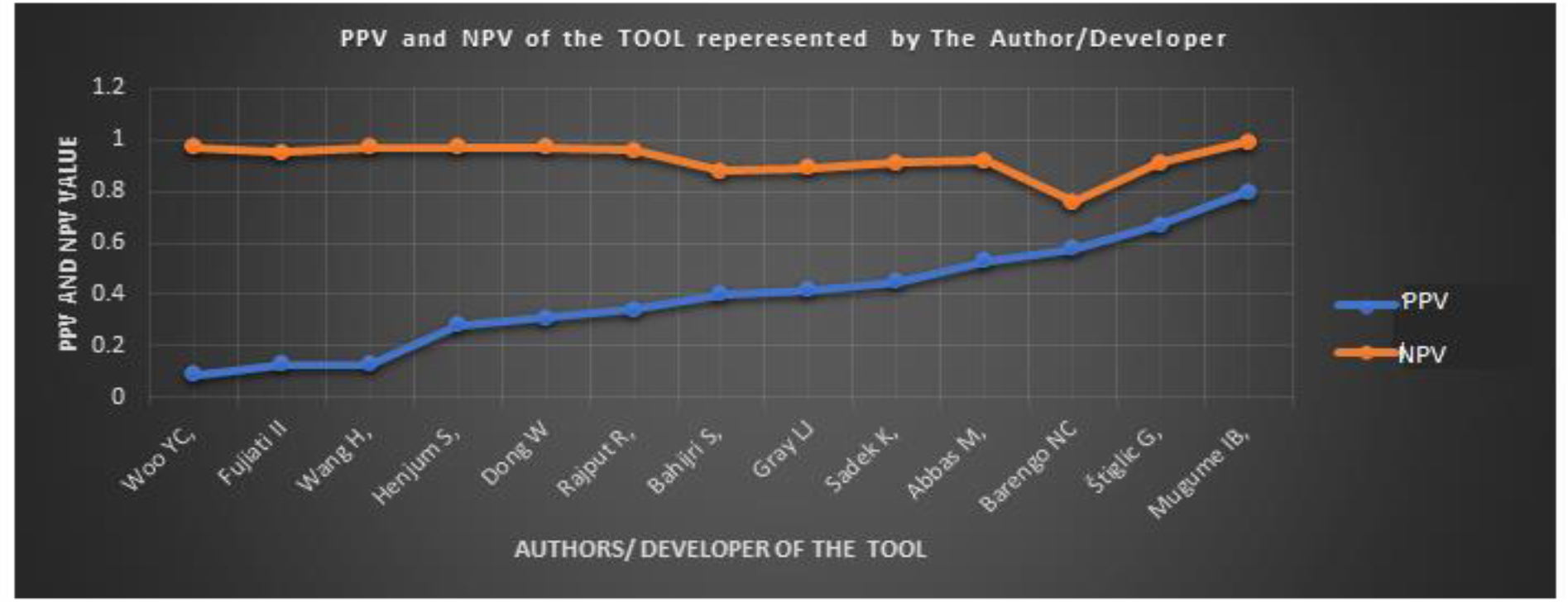
The predictor Tool PPV and NPV in reference to the Authors

**Figure 11.**
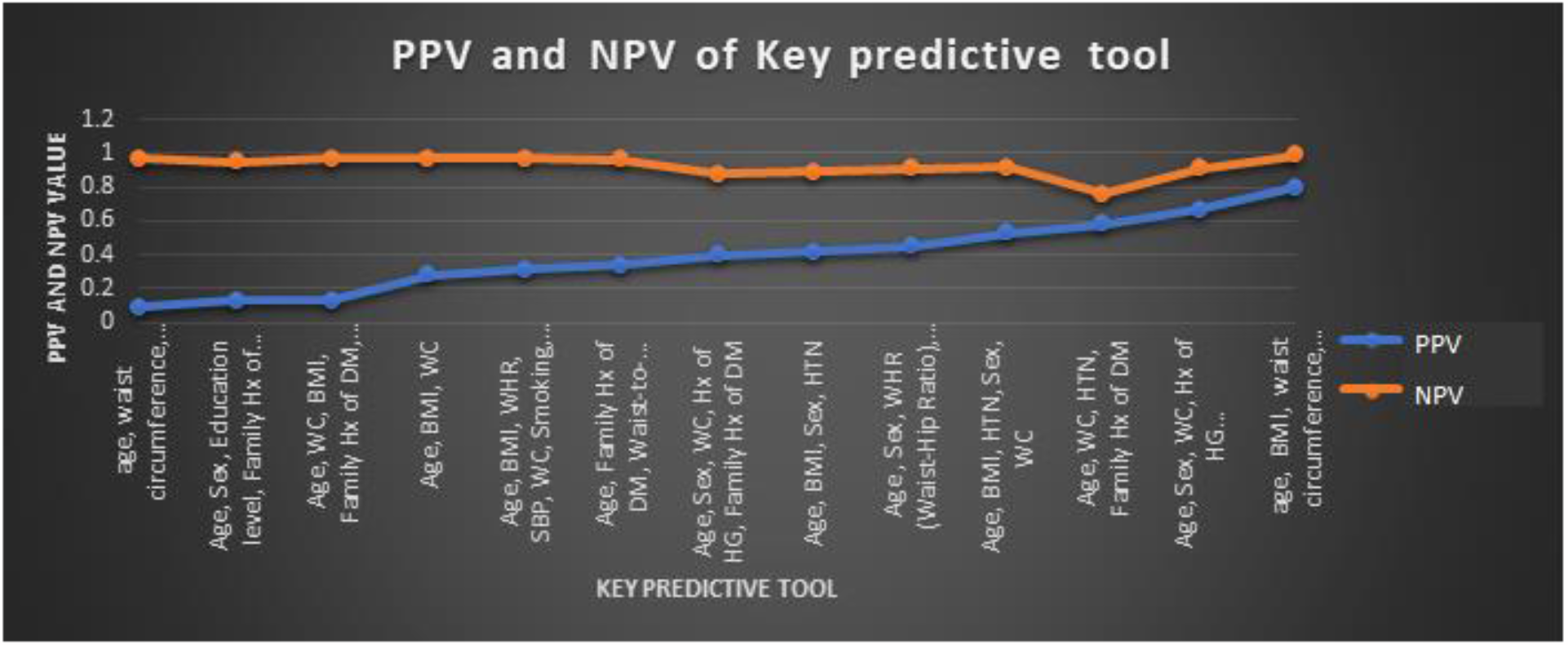
The PPV and NPV of Key predictive tool

## Discussion

Diabetes noninvasive Screening tests have the potential to identify prediabetes at an early stage and, thus, a more treatable abnormality, as a result, saves lives.^27^ This review identified 24 risk key prediction parameters to develop the tools that used only non-laboratory risk predictors to detect individuals with pre-DM from 22 published studies conducted from 2013 to 2023. The tools included similar risk predictors in 60% to 90%: age, hypertension, family history of DM, and obesity (represented by BMI and WC), regardless of the origin of the Article’s geographic location. The Tools had promising discrimination ability in case finding of pre-DM as shown by its Sensitivity/Specificity and PPV/NPV.^27^

Sensitivity and specificity indicate the Tool’s effectiveness to the ‘gold standard’ referent.^27,28^ So, sensitivity and specificity indicate the concordance of a Tool test for a chosen ‘gold standard’ reference, in these studies, FBS, RBS, 1 h PG, 2h PG, HbA1C. It was noted that some reporting variation was found among the studies on cutting points for pre-DM diagnosis. This variation may originate from the Prediabetes and Diabetes definition source. It is wise to bear this fact in mind when interpreting the discrepancy of the result since this, in turn, affects the sensitivity/specificity of the developed Tool.

As shown in Figures 7,8,9,10 and 11, there are noticeable tradeoffs between sensitivity and specificity and between positive predictive values (PPVs) and negative predictive values (NPVs).^27^ A highly value-sensitive test means that there are few false negative results, and thus, fewer disease cases are missed. The specificity of a test is its ability to detect an individual who does not have a disease as a negative result.^27,28^ A high specificity test means that there are few false positive results. For example, the Tool developed by Tan and his colleague revealed a sensitivity/Specificity of 0.92 (92%)/0.66(66%).^20^ (see Figures 7 and 8) The Tool, according to the article result, only 8% of cases are missed, and 44% are falsely included as a positive.

Meanwhile, PPV and NPV assess the people being screened.^27,28^ The value of PPV/NPV indicates the likelihood that the Tool can successfully identify whether people have prediabetes or not. PPV answers: what is a person’s likelihood of true positive if the screening tool yields a positive result? Or what is the probability that a person has Prediabetes? On the other hand, NPV gives us insight: if the screening tool yields a negative result, what is the probability that the person does not have Prediabetes?^27,28^ For instance, the study conducted by Mugume IB and his colleague^17^ (Figures 10 and 11) revealed that the Tool they developed has 0.80 (80%) PPV and 0.99 (99%) NPV. That means the likelihood or probability of the person screened by this Tool being positive for Prediabetes, being true Prediabetes is 80%, and falsely labeling the person as having Pre-DM is 20%. If the Tool shows a negative result (i.e., Free of Prediabetes), its probability of being truly free of Prediabetes is 99% and misses only 1%. So, the Tool developed by these researchers looks promising for our goal of screening Diabetes by noninvasive method to answer this review research question. To optimize the screening procedure capacity, it is essential to identify the target population likely to benefit most, considering the prevalence of prediabetes, such as age, gender, race, Obesity, known medical condition, or another risk factor (e.g., family history of diabetes).^27^

Hence, the knowledge gained by screening using a tool like this can increase health awareness and motivate the person to develop intervention strategies to prevent the occurrence of Diabetes, for instance, by modifying lifestyle. Modifying parameters indicated in this review, such as number of hours of sleep, Exercise (physical activity), and obesity [represented by BMI, WC, and waist-to-hip ratio (WHR) can contribute to the prevention strategies of Diabetes.^1,2^

## Conclusions

This review revealed potential Tools to screen Diabetes noninvasively and non-laboratory methods, which can be applied individually or population-based screening purposes where the prevalence of the disorder is high regardless of involving health professionals. The findings promise to make screening tools easily accessible and applicable. In the review, the most common risk predictors were Age, WC, Hypertension, BMI, and Family history of Diabetes, which needs considerable attention in the planning process of prevention. (see figure 3) Early screening and intervention by developing healthy lifestyles can contribute to successfully preventing and controlling Diabetes.^1,2^

The strength of this review is that it included an up-to-date synthesis of the results from studies conducted in 13 countries and different continents (which addresses the cultural and race diversity) of the world, conducted in the last ten years. The review result explicitly showed the value of sensitivity, Specificity, PPV, and NPV of the developed Tool to compare and choose from to facilitate the case finding (pre-DM).^27,28^ However, several limitations regarding these Scoping reviews must be acknowledged. The bias assessment of the included Article is not performed, which can be addressed in the future by a systematic review and meta-analysis when time constraint is not an issue.^27^ In addition, the inclusion of only studies published in the English language could miss some pertinent tools from studies written in another language.

A systematic meta-analysis and RCT on the Tool described in this scoping review is recommend to identify any bias during the development of the Tool and possible generalizability of the best Tool for worldwide applicability.

## Data Availability

All data produced in the present work are contained in the manuscript.

